# Visible and real dynamics of the COVID-19 pandemic in Ukraine in the spring of 2021

**DOI:** 10.1101/2021.06.13.21258838

**Authors:** Igor Nesteruk

## Abstract

The visible and real sizes the last COVID-19 epidemic wave in Ukraine, estimated in March 2021 with the use of generalized SIR-model, are compared with number of cases registered in the spring of 2021. We have used the optimal value of the visibility coefficient in order to estimate the real numbers of accumulated cases, real daily numbers of new cases and real number of infectious persons. The results show that the latest prediction for Ukraine is in rather good agreement with observations, but the daily number of new cases decreases more slowly than theoretical predictions. The large real number of infectious people threatens the emergence of new strains of coronavirus and the beginning of new epidemic waves.

## Introduction

The studies of the COVID-19 pandemic dynamics are complicated by a very large percentage of asymptomatic patients who are not registered by medical statistics. For Ukraine, different simulation and comparison methods were based on official accumulated number of laboratory-confirmed cases [1, 2] (these figures coincided with the official WHO data sets [3], but WHO stopped to provide the daily information in August 2020). In particular, the classical SIR model [4-6] was used in [7, 8]. The weakening of quarantine restrictions, changes in the social behavior and the coronavirus mutations caused changes in the epidemic dynamics and corresponding parameters of models. To detect and simulate these new epidemic waves, a simple method of numerical differentiations of the smoothed number of cases and generalized SIR-model were proposed and used in [8-10]. In particular, nine epidemic waves in Ukraine were calculated [8-10]. Since the number of laboratory-confirmed COVID-19 cases is much higher than the real one [11-16], the identification algorithm for SIR-parameters [17] was modified in [18, 19] in order to assess the extent of data incompleteness and determine the true sizes of the COVID-19 epidemic in Ukraine and in Qatar [20]. In this paper we will compare the predictions proposed in March 2021 in preprint [18] with the official pandemic dynamics in Ukraine in March-June 2021. The visible and real dimensions of the pandemic in Ukraine will be estimated and discussed.

## Data

We will use the data set regarding the accumulated numbers of laboratory-confirmed COVID-19 cases in Ukraine from national sources [1, 2]. The corresponding numbers *V*_*j*_ and moments of time *t*_*j*_ (measured in days) are shown in Table 1. The values *V*_*j*_ for the period March 11-24, 2021 have been used in [18,19] for SIR simulations of the tenth epidemic wave in Ukraine. Other values will be used to control the accuracy of predictions and pandemic dynamics.

**Table 1.** **Cumulative numbers of laboratory-confirmed Covid-19 cases in Ukraine *V*_*j*_ according to the national statistics, [1, 2].**

| Day in March 2021 | Number of cases, $V_j$ | Day in April 2021 | Number of cases, $V_j$ | Day in May 2021 | Number of cases, $V_j$ | Day in June 2021 | Number of cases, $V_j$ |
| --- | --- | --- | --- | --- | --- | --- | --- |
| 1 | 1357470 | 1 | 1711630 | 1 | 2083180 | 1 | 2206836 |
| 2 | 1364705 | 2 | 1731971 | 2 | 2085938 | 2 | 2209417 |
| 3 | 1374762 | 3 | 1745709 | 3 | 2088410 | 3 | 2211683 |
| 4 | 1384917 | 4 | 1755888 | 4 | 2090986 | 4 | 2213580 |
| 5 | 1394061 | 5 | 1769164 | 5 | 2097024 | 5 | 2214517 |
| 6 | 1401228 | 6 | 1784579 | 6 | 2105428 | 6 | 2215052 |
| 7 | 1406800 | 7 | 1803998 | 7 | 2114138 | 7 | 2216654 |
| 8 | 1410061 | 8 | 1823674 | 8 | 2119510 | 8 | 2218039 |
| 9 | 1416438 | 9 | 1841137 | 9 | 2122327 | 9 | 2219824 |
| 10 | 1425522 | 10 | 1853249 | 10 | 2124535 | - | - |
| 11 | 1438468 | 11 | 1861105 | 11 | 2129073 |  |  |
| 12 | 1451744 | 12 | 1872785 | 12 | 2135886 |  |  |
| 13 | 1460756 | 13 | 1887338 | 13 | 2143448 |  |  |
| 14 | 1467548 | 14 | 1903765 | 14 | 2150244 |  |  |
| 15 | 1477190 | 15 | 1921244 | 15 | 2153864 |  |  |
| 16 | 1489023 | 16 | 1936228 | 16 | 2156000 |  |  |
| 17 | 1504076 | 17 | 1946510 | 17 | 2160095 |  |  |
| 18 | 1519926 | 18 | 1953016 | 18 | 2165233 |  |  |
| 19 | 1535218 | 19 | 1961956 | 19 | 2170398 |  |  |
| 20 | 1546363 | 20 | 1974118 | 20 | 2175382 |  |  |
| 21 | 1554256 | 21 | 1990353 | 21 | 2179988 |  |  |
| 22 | 1565732 | 22 | 2004630 | 22 | 2182521 |  |  |
| 23 | 1579906 | 23 | 2017341 | 23 | 2183855 |  |  |
| 24 | 1596575 | 24 | 2025271 | 24 | 2186463 |  |  |
| 25 | 1614707 | 25 | 2030333 | 25 | 2189858 |  |  |
| 26 | 1632131 | 26 | 2038248 | 26 | 2193367 |  |  |
| 27 | 1644063 | 27 | 2047838 | 27 | 2196673 |  |  |
| 28 | 1652409 | 28 | 2059465 | 28 | 2199769 |  |  |
| 29 | 1662942 | 29 | 2069537 | 29 | 2201472 |  |  |
| 30 | 1674168 | 30 | 2078086 | 30 | 2202494 |  |  |
| 31 | 1691737 | - | - | 31 | 2204631 |  |  |

### Generalized SIR model and parameter identification procedure

In [18, 19] the generalized SIR-model and the exact solution of the set of non-linear differential equations relating the number of susceptible *S*, infectious *I* and removed persons *R* was used (see, e.g., [21]). This solution uses the function

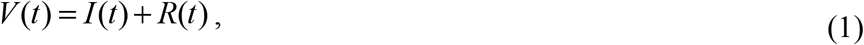

corresponding to the number of victims or the cumulative confirmed number of cases. Its derivative:

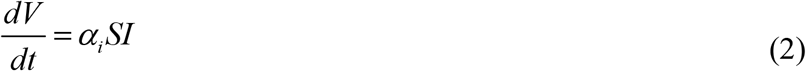

yields the estimation of the average daily number of new cases. When the registered number of victims *V*_*j*_ is a random realization of its theoretical dependence (1), the exact solution presented in [21] depends on five parameters (*α*_*i*_ is one of them). The details of the optimization procedure for their identification can be found in [17].

If we assume, that data set *V*_*j*_ is incomplete and there is a constant coefficient *β*_*i*_ ≥ 1, relating the registered and real number of cases during the *i-th* epidemic wave:

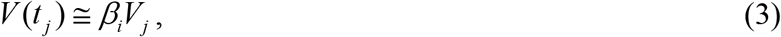

the number of unknown parameters increases by one. The values *V*_*j*_, corresponding to the moments of time *t*_*j*_ from the period March 11-24, 2021 have been used in [18,19] to find the optimal values of these parameters corresponding to the tenth epidemic wave in Ukraine. In particular, the optimal value of the visibility coefficient *β*_10_ =3.7 was calculated.

### Monitoring changes in epidemic parameters and selection of epidemic waves

Changes in the epidemic conditions (in particular, the peculiarities of quarantine and its violation, situations with testing and isolation of patients, emergence of new strains of the pathogen) cause the changes in its dynamics, i.e. so known epidemic waves. To control these changes we can use daily or weekly numbers of new cases and their derivatives (see, e.g., [8, 10, 22]). Since these values are random, we need some smoothing (especially for daily amounts, which are also characterized by some weekly periodicity). For example, we can use the smoothed daily number of accumulated cases:

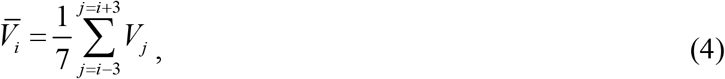

The first and second derivatives can be estimated with the use of following formulas:

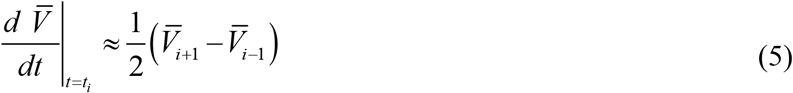

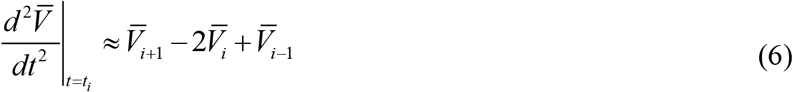

Fig. 1 illustrates the application of formulae (4)-(6) for the COVID-19 pandemic dynamics in Ukraine in the spring and the summer of 2020, [10].

**Fig. 1.**
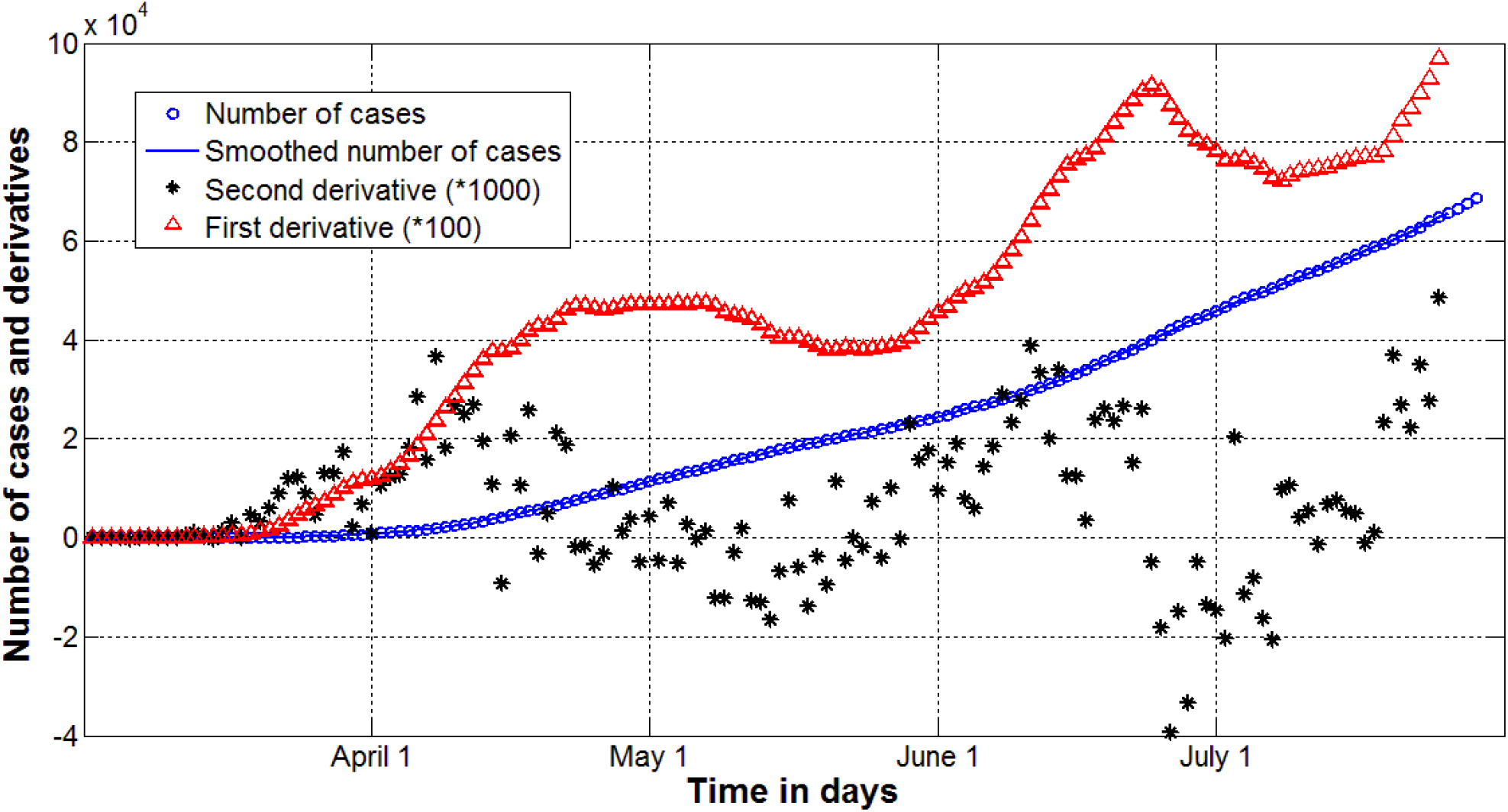
Epidemic dynamics in Ukraine in the spring and the summer of 2020, [10]. Accumulated number of cases (blue markers and line, eq. (4)). Red markers represent first derivative (eq. (5)) multiplied by 100, black one show the second derivative (eq. (6)) multiplied by 1000.

## Results and Discussion

The results of SIR simulation performed in [18, 19] are shown in Fig. 2. Blue lines represent the estimations of the real sizes of the tenth COVID-19 in Ukraine for the optimal value *β*_10_ =3.7 in relationship (3): *V(t)=I(t)+R(t)* – solid; dashed one represents the number of infectious persons multiplied by 5, i.e. *I* (*t*) ×5 ; dotted line shows the derivative (*dV* / *dt*)×100 calculated with the use of formula (2). Black solid line shows the “visible” dependence *V(t)=I(t)+R(t)* calculated in [18,19] with the use of assumption that *β*_10_ =1. Red “circles” and “stars” correspond to the accumulated numbers of cases registered during the period of time taken for SIR simulations (March 11-24, 2021) and beyond this time period, respectively.

**Fig. 2.**
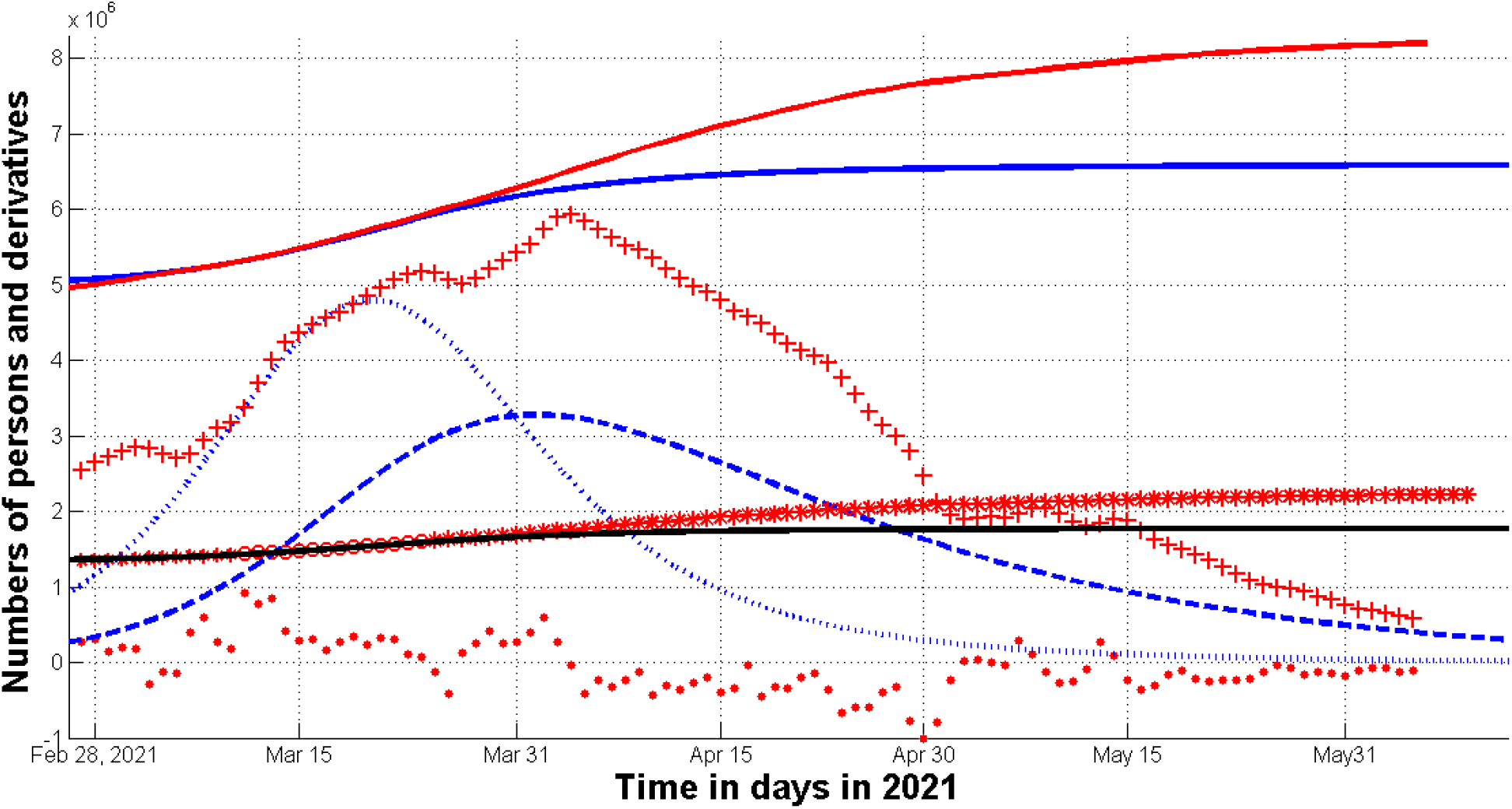
Real and visible COVID-19 epidemic dynamics in Ukraine in the spring of 2021. The results of SIR simulations of the tenth wave at the optimal value *β*_10_ =3.7 are shown by blue lines, [18, 19]. Numbers of victims *V(t)=I(t)+R(t)* – solid lines; numbers of infected and spreading *I(t)* multiplied by 5 – dashed; derivatives *dV/dt* (eq. (2)) multiplied by 100 – dotted. The black solid line shows the “visible” dependence *V(t)=I(t)+R(t)* calculated with the use of assumption that *β*_10_ =1, [18, 19]. The red solid line shows smoothed accumulated number of laboratory-confirmed cases (eq. (4)) multiplied by the optimal value *β*_10_ =3.7. Red “circles” and “stars” correspond to the accumulated numbers of cases registered during the period of time taken for SIR simulations (March 11-24, 2021) and beyond this time period, respectively. The red “crosses” and “dots” show the first derivative (5) multiplied by 100*β*_10_ and the second derivative (6) multiplied by 1000, respectively.

To illustrate the changes in the epidemic dynamics, the derivatives (5) and (6) were calculated with the use of *V*_*j*_ values presented in Table 1. The red “crosses” and “dots” show the first derivative (5) multiplied by 100*β*_10_ and the second derivative (6) multiplied by 1000, respectively. Fig. 2 shows minima of the smoothed daily number of new cases (eq. (5)) in March and May 2021 (see red “crosses”) and corresponding jumps of second derivative (6) (see red “dots”). These facts illustrate the changes in the dynamics which have to be simulated by segmentation of new epidemic waves and corresponding new SIR simulations for each one. The derivative (2) cannot have any minimum for a fixed epidemic wave. There is only one maximum, corresponding to zero point of *d*^*2*^*V/dt*^*2*^ (see [21] and the blue dotted line in Fig. 2).

The reason for changes in the typical epidemic dynamics could be changes in social behavior (especially in the first half of May, during the celebration of Orthodox Easter and May holidays) and with the emergence of new strains of coronavirus. Therefore, it is difficult to expect very good accuracy from SIR simulations based on the data from the period March 11-24, 2021. During April and May 2021, the optimal values of the model parameters (including the value of the visibility coefficient *β*) could change. However, the difference between the theoretical curve *V(t)=I(t)+R(t)* (see the blue solid line in Fig. 2) and the accumulated laboratory-confirmed number of cases (smoothed according to formula (4) and multiplied by the value *β*_10_ =3.7 according to formula (3), see the red solid line in Fig. 2) does not exceed 20%. Accuracy can be increased by selecting individual epidemic waves and calculating new optimal values of parameters (including values *β*_11_, *β*_12_, etc.).

Unfortunately, mathematical modeling can hardly answer the question of whether we will have new waves of the epidemic after June 2021. Because even strict quarantine measures does not prevent the emergence of new strains of coronavirus both domestically and imported from abroad. The number of infectious persons in Ukraine is still very high. According to the optimistic estimation (shown by the blue dashed line in Fig. 2), the number of infectious person will be around 40,000 in mid-June 2021. This fact and the delays in vaccination make the probability of the emergence of new domestic strains of coronavirus very high. In addition, the unfavorable course of the pandemic in India [23, 24] and other countries increases the likelihood of importing new strains to Ukraine.

Compliance with social distance and other quarantine measures remains very important. Mass violations of simple rules last summer led to new waves of the epidemic (see Fig. 1). Since the real stabilization of the epidemic dynamics occurred later than in the theoretical prediction (compare red and blue solid lines in Fig. 2), the epidemic duration may increase. Even in the case of an optimistic scenario (without the emergence of new waves), the end of the epidemic in Ukraine should not be expected earlier than August 2022 (see [18, 19]). The government should closely monitor the average daily number of cases (which can be calculated by formulas (4) and (5)) and immediately intensify quarantine measures in case of its increase. As of June 10, 2021, there is no cause for concern (red crosses in Fig. 2 show a downward trend in the derivative (5)).

## Data Availability

data is in the text

## Acknowledgements

The author is grateful to Oleksii Rodionov for his help in collecting and processing data.

